# SARS-CoV-2 Convalescent Sera Binding and Neutralizing Antibody Concentrations Compared with COVID-19 Vaccine Efficacy Estimates Against Symptomatic Infection

**DOI:** 10.1101/2021.11.24.21266812

**Authors:** Amy J. Schuh, Panayampalli S. Satheshkumar, Stephanie Dietz, Lara Bull-Otterson, Myrna Charles, Chris Edens, Jefferson M. Jones, Kristina L. Bajema, Kristie E.N. Clarke, L. Clifford McDonald, Sadhna Patel, Kendra Cuffe, Natalie J. Thornburg, Jarad Schiffer, Kelly Chun, Monique Bastidas, Manory Fernando, Christos J. Petropoulos, Terri Wrin, Suqin Cai, Dot Adcock, Deborah Sesok-Pizzini, Stanley Letovsky, Alicia M. Fry, Aron J. Hall, Adi V. Gundlapalli

## Abstract

Previous vaccine efficacy (VE) studies have estimated neutralizing and binding antibody concentrations that correlate with protection from symptomatic infection; how these estimates compare to those generated in response to SARS-CoV-2 infection is unclear. Here, we assessed quantitative neutralizing and binding antibody concentrations using standardized SARS-CoV-2 assays on 3,067 serum specimens collected during July 27, 2020-August 27, 2020 from COVID-19 unvaccinated persons with detectable anti-SARS-CoV-2 antibodies using qualitative antibody assays. Quantitative neutralizing and binding antibody concentrations were strongly positively correlated (r=0.76, p<0.0001) and were noted to be several fold lower in the unvaccinated study population as compared to published data on concentrations noted 28 days post-vaccination. In this convenience sample, ∼88% of neutralizing and ∼63-86% of binding antibody concentrations met or exceeded concentrations associated with 70% COVID-19 VE against symptomatic infection from published VE studies; ∼30% of neutralizing and 1-14% of binding antibody concentrations met or exceeded concentrations associated with 90% COVID-19 VE. These data support observations of infection-induced immunity and current recommendations for vaccination post infection to maximize protection against symptomatic COVID-19.

## BACKGROUND

As of October 18, 2021, 66.7% of the US population 12 years of age and older has been fully vaccinated for COVID-19 [1]. A recent Gallup survey found that 18% of Americans would not agree to be vaccinated if a US Food and Drug Administration (FDA)-approved COVID-19 vaccine were available to them immediately at no cost [2]. One of the primary reasons cited for vaccine hesitancy was a history of SARS-CoV-2 infection and resultant antibodies.

Cumulative evidence indicates that SARS-CoV-2 antibodies are protective against SARS-CoV-2 re-infection [3]. A series of non-human primate challenge studies demonstrated the central role of SARS-CoV-2 neutralizing antibodies in protection from re-infection [4-6]. A randomized clinical trial involving the subcutaneous administration of REGEN-COV, a combination of two SARS-CoV-2 neutralizing monoclonal antibodies, or placebo within 96 hours of SARS-CoV-2 exposure demonstrated that REGEN-COV prevented symptomatic COVID-19 and asymptomatic SARS-CoV-2 infection [7]. A longitudinal study of >12,000 health care workers showed that SARS-CoV-2 infection-induced protective immunity lasts for at least 6 months [8].

The establishment of the First World Health Organization (WHO) International Standard for Anti-SARS-CoV-2 Immunoglobulin [9] for quantitative assessment of neutralizing and binding antibody concentrations has made it possible for COVID-19 vaccine efficacy (VE) studies [10-13] to describe and propose standardized immune correlates of protection against symptomatic infection (or risk of symptomatic infection) in fully-vaccinated persons (i.e., 2 weeks after their second dose in a 2-dose series, such as Pfizer-BioNTech (BNT162b2), Moderna (mRNA-1273), or AstraZeneca (ChAdOx1) vaccines OR 2 weeks after a single-dose vaccine, such as Johnson & Johnson’s Janssen [JNJ-78436735] vaccine) across vaccine trials that have used different antibody assays. Comparing antibody concentrations of unvaccinated persons with anti-SARS-CoV-2 antibodies to COVID-19 vaccinated cohorts and to estimated antibody concentrations associated with COVID-19 vaccine effectiveness would accomplish three goals: 1) improve understanding of the population distribution of antibody concentrations in response to infection and vaccination, 2) help establish relationship of quantitative antibody levels to those associated with protection against illness from published studies, and 3) inform better targeted messaging for universal COVID-19 vaccination.

Numerous SARS-CoV-2 serological assays have been developed over the course of the COVID-19 pandemic to measure virus-specific antibody responses [14]. Persons who recover from SARS-CoV-2 infection or receive a COVID-19 vaccine typically develop virus-specific neutralizing antibodies, with most of these antibodies directed against the immunodominant receptor binding domain (RBD) of the spike (S) protein [15, 16]. Efficacy trials of the ChAdOx1 [11] and mRNA-1273 [12] vaccines showed that higher anti-SARS-CoV-2 S IgG, anti-SARS-CoV-2 RBD IgG, and SARS-CoV-2 neutralizing antibody concentrations were correlated with a reduced risk of symptomatic infection. Both trials determined antibody concentrations associated with varying levels of VE against symptomatic COVID-19 [11, 12]. The ChAdOx1 vaccine trial estimated that 70% and 90% VE against symptomatic COVID-19 was associated with 50% neutralizing antibody titer (NT_50_) concentrations of 3.7 and 64.1 international units per mL (IU/mL), respectively, and 70% and 90% VE was associated with anti-RBD IgG antibodies concentrations of 165.0 and 2360.0 binding antibody units per mL (BAU/mL), respectively [11]. The mRNA-1273 vaccine trial estimated that 70% and 90% VE against symptomatic COVID-19 was associated with NT_50_ concentrations of 4.0 and 83.0 IU/mL, respectively, and 70% and 90% VE was associated with anti-RBD IgG antibodies concentrations of 8.0 and 775.0 BAU/mL, respectively [12]. However, it remains unclear how these antibody concentrations compare to those generated in response to SARS-CoV-2 infection in COVID-19 unvaccinated persons. To answer this question, we tested 3,067 pre-vaccination sera collected as part of a nationwide commercial laboratory seroprevalence study led by the US Centers for Disease Control and Prevention (CDC) [17] that had previously tested positive for anti-SARS-CoV-2 antibodies using standardized anti-SARS-CoV-2 RBD IgG and SARS-CoV-2 pseudovirus neutralization antibody assays, and then compared these antibody concentrations to concentrations measured in COVID-19 vaccinated cohorts (28 days post-vaccination) and to those associated with 70% and 90% COVID-19 VE against symptomatic infection for two of the vaccines for which such data are published to date.

## METHODS

### Specimen Source and Study Design

Sera were collected by two US-based commercial laboratories for routine or acute clinical care (e.g., cholesterol screening or a sick visit) as part of a nationwide seroprevalence study; detailed methods have previously been described [17]. Briefly, blood specimens collected for COVID-19-related reasons were excluded. Sera were tested [17] for anti-SARS-CoV-2 antibodies using one of the following three qualitative assays issued emergency use authorization by the FDA that were in use in the clinical laboratories: 1) Architect™ SARS-CoV-2 IgG Assay (nucleocapsid (N) protein; Abbott, Chicago, IL), 2) VITROS® Anti-SARS-CoV-2 IgG Assay (S protein; Ortho-Clinical Diagnostics, Raritan, NJ), and 3) Elecsys® Anti-SARS-CoV-2 Assay (N protein; Roche, Indianapolis, IN) (Supplementary figure 3). Among 84,683 total serum samples collected during July 27, 2020-August 27, 2020 identified as anti-SARS-CoV-2 antibody positive and had linked age and sex information (as part of the larger serosurveillance study) [17], 3067 serum specimens were selected by convenience sampling for reflex testing with anti-SARS-CoV-2 quantitative IgG and neutralizing antibody assays. Data on race, ethnicity, and occurrence or date of SARS-CoV-2 infection or symptoms were not available; data on prior SARS-CoV-2 quantitative reverse transcription polymerase chain reaction qRT-PCR test results were not available for most persons.

### Ethics

This activity was reviewed by CDC and was conducted consistent with applicable federal law and CDC policy (45 C.F.R. part 46, 21 C.F.R. part 56; 42 U.S.C. Sect. 241(d); 5 U.S.C. Sect. 552a; 44 U.S.C. Sect. 3501 et seq.). Informed consent was waived, as all data were deidentified.

### PhenoSense® CoV Neutralizing Antibody Assay

Neutralizing antibodies against the SARS-CoV-2 S protein were measured using the PhenoSense CoV Neutralizing Antibody Assay® [18]. To measure SARS-CoV-2 neutralizing antibodies, human immunodeficiency virus-1 pseudovirions expressing the SARS-CoV-2 S protein were prepared by co-transfecting HEK293 producer cells with an HIV-1 genomic vector and a SARS-CoV-2 envelope protein expression vector. Serial dilutions of sera were incubated with recombinant pseudovirions and neutralizing activity was assessed by measuring the inhibition of luciferase activity in HEK293 target cells co-expressing the angiotensin-converting enzyme 2 and transmembrane serine protease 2 receptors. Fifty percent neutralizing antibody titers (NT_50_) were expressed as the reciprocal of the serum dilution conferring 50% inhibition of pseudovirus infection. Calibration of the PhenoSense CoV Neutralizing Antibody Assay® with the First WHO International Standard for Anti-SARS-CoV-2 Immunoglobulin (20/136, National Institute for Biological Standards and Controls, UK) [9] allowed us to generate a calibration factor of 0.0653 (S protein containing G614) and convert NT_50_ values from titers to international units per mL (IU/mL) by multiplying by the calibration factor.

### Cov2Quant IgG® Assay

Anti-SARS-CoV-2 RBD IgG in sera were quantified using the electrochemiluminescent (ECL) Cov2Quant IgG® Assay [19]. Briefly, sera were incubated with biotin-conjugated, recombinant SARS-CoV-2 RBD antigen bound to a streptavidin, carbon-coated microtiter well plate. After washing the plate, the wells were incubated with ruthenium-conjugated anti-IgG and a final wash was performed to remove unbound material. Voltage was applied to the metallic electrode on the base of the plate and a photomultiplier was used to measure the ECL signal generated from each well. The signal in relative light units (RLU) is directly proportional to the anti-RBD IgG concentration in sera. Each assay batch included a dilution series of affinity-purified human IgG standard that was used to calculate the anti-SARS-CoV-2 RBD IgG concentration of each serum. Calibration of the Cov2Quant IgG® Assay with the WHO International Standard allowed us to generate a calibration factor of 25 and convert anti-SARS-CoV-2 RBD IgG concentrations from µg/mL to binding antibody units per mL (BAU/mL) by multiplying by the calibration factor.

### Comparison of Study Antibody Data to Antibody Concentrations Associated with COVID-19 VE

Of the COVID-19 vaccines in use around the world currently, serological correlates of protection thresholds associated with VE have been estimated for ChAdOx1 [11] and mRNA-1273 [12] using the First WHO International Standard for Anti-SARS-CoV-2 Immunoglobulin. As the quantitative antibody assays used in our study were calibrated to the same standard, we were able to compare antibody concentrations in our study to those estimated to be associated with 70% and 90% COVID-19 VE to ChAdOx1 and mRNA-1273.

### Statistical Analyses

Data management tasks, and statistical analyses were performed using SAS version 9.4 (SAS Institute Inc., Cary, NC) and GraphPad Prism 9.0.0 (GraphPad Software, San Diego, CA). T-tests on log_10_-transformed data were used to determine if there were statistically significant differences between geometric mean anti-SARS-CoV-2 RBD IgG and NT_50_ concentrations by sex. ANOVAs on log_10_-transformed data were used to determine if there were statistically significant differences between geometric mean anti-SARS-CoV-2 RBD IgG and NT_50_ concentration according to previous qualitative antibody test and age category. Post-hoc Tukey’s tests were performed to identify previous qualitative antibody tests and age categories with significantly different anti-SARS-CoV-2 RBD IgG and NT_50_ concentrations; p-values were adjusted to account for multiple comparisons. Pearson’s correlation was used to assess the overall relationship between log_10_anti-RBD IgG and log_10_NT_50_ concentrations, as well as the relationships between the two variables across sex and age category. Two-sided p-values <0.05 were considered statistically significant.

## RESULTS

Of the overall convenience sample of 3,067 serum specimens collected during July 27, 2020-August 27, 2020 with detectable anti-SARS-CoV-2 antibodies on a qualitative assay, 1,309 were from males (42.7%) and 1,758 (57.3%) were from females, 779 (25.4%) were from persons aged <18 years, 982 (32.0%) were from persons aged 18–49 years, 798 (26.0%) were from persons aged 50–64 years, and 508 (16.6%) were from persons aged ≥65 years.

Most of the serum specimens (n=2568, 83.7%) tested positive for both quantitative SARS-CoV-2 neutralizing antibodies and quantitative anti-SARS-CoV-2 RBD IgG; 271 sera (8.8%) tested negative for both neutralizing antibodies and anti-RBD IgG, 83 sera (2.7%) tested negative for neutralizing antibodies only, and 145 sera (4.7%) tested negative for anti-RBD IgG only. Geometric mean NT_50_ concentrations differed significantly according to the qualitative antibody test type that was used to screen sera for study inclusion (p<0.0001; Supplementary Figure 4A), while geometric mean anti-RBD IgG concentrations did not differ significantly according to the qualitative antibody test type that was used to screen sera for study inclusion (p=0.02998; Supplementary Figure 4B).

The overall geometric mean NT_50_ concentration was 28.8 IU/mL (95% confidence interval (CI): 26.8, 31.0) and did not differ significantly according to sex (male: 30.9 IU/mL, 95% CI: 27.4, 34.8; female: 27.3 IU/mL, 95% CI: 25.0, 30.0; p=0.1088) but differed significantly according to age category (<18 years: 31.3 IU/mL, 95% CI: 28.4, 34.6; 18–49 years: 23.1 IU/mL, 95% CI: 20.3, 26.2; 50–64 years: 38.0 IU/mL, 95% CI: 32.5, 44.4; ≥65 years: 25.0 IU/mL, 95% CI: 20, 31.3; p<0.0001; Figure 1A). Persons aged <18 years had significantly higher NT_50_ concentrations compared to persons aged 18– 49 years (p=0.0108), and persons aged 50–64 years had significantly higher NT_50_ concentrations compared to persons aged 18–49 years (p<0.0001) and persons aged ≥65 years (p=0.0020) (Figure 1A). The overall geometric mean anti-RBD IgG concentration was 162.5 BAU/mL (95% CI: 152.9, 172.7) and did not differ significantly according to sex (male: 171.3 BAU/mL, 95% CI: 155.6, 188.5; female: 156.3 BAU/mL, 95% CI: 144.4, 169.1; p=0.1447) but did differ significantly according to age category (<18 years: 218.2 BAU/mL, 95% CI: 199.2, 239.1; 18–49 years: 122.2 BAU/mL, 95% CI: 109.8, 136.1; 50–64 years: 189.0 BAU/mL, 95% CI: 167.2, 213.7; ≥65 years: 141.4 BAU/mL, 95% CI: 117.8, 169.8; p<0.0001; Figure 1B). Like the age-associated patterns of neutralizing antibody concentrations, persons aged <18 years had significantly higher anti-RBD IgG concentrations compared to persons aged 18–49 years (p<0.0001) and persons aged ≥65 years (p<0.0001), and persons aged 50–64 years had significantly higher anti-RBD IgG concentrations compared to persons aged 18–49 years (p<0.0001) and persons aged ≥65 years (p=0.0147) (Figure 1B).

**Figure 1.**
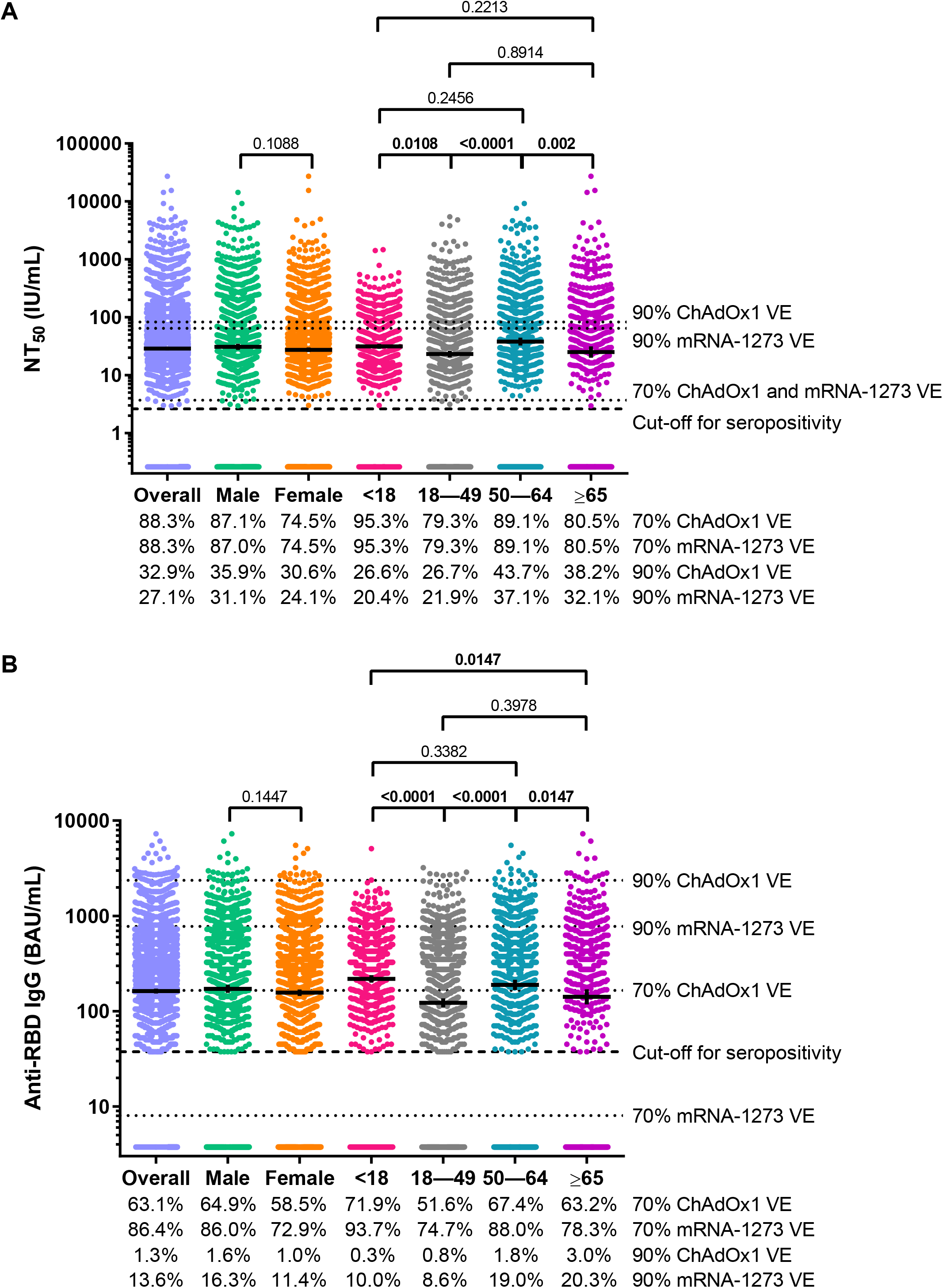
SARS-CoV-2 50% neutralizing antibody titer (NT_50_) concentrations and anti-SARS-CoV-2 receptor binding domain (RBD) IgG concentrations for the overall convenience sample of 3,067 serum specimens collected during July 27,2020-August 27, 2020 with detectable anti-SARS-CoV-2 antibodies on a qualitative assay, and according to sex and age category. A) SARS-CoV-2 NT_50_ concentrations in international units per mL (IU/mL) and B) Anti-SARS-CoV-2 RBD IgG concentrations in binding antibody units per mL (BAU/mL). Horizontal bars represent geometric means, vertical error bars represent 95% confidence intervals, and dashed horizontal lines represent assay cut-off values for seropositivity. P-values from t-tests (sex) and post-hoc Tukey tests (age class) are shown for each sex and age class comparison. Bolded p-values denote statistical significance (p<0.05). Dotted horizontal lines represent antibody concentrations associated with 70% and 90% ChAdOx1 [11] and mRNA-1273 [12] vaccine efficacy (VE). The percentage of sera with antibody concentrations that meet or exceed the concentrations represented by each of the horizontal dotted lines are shown below the charts.

We found an overall significant, strong positive correlation between NT_50_ concentrations and anti-RBD IgG concentrations (r=0.76, p<0.0001; Figure 1A). The strength of the relationship remained consistent across sex (male: r=0.76, p<0.0001; female: r=0.76, p<0.0001; Figure 1B) and increased with increasing age category (<18 years: r=0.67, p<0.0001; 18–49 years: r=0.69, p<0.0001; 50–64 years: r=0.79, p<0.0001; ≥65 years: r=0.88, p<0.0001; Figure 2).

**Figure 2.**
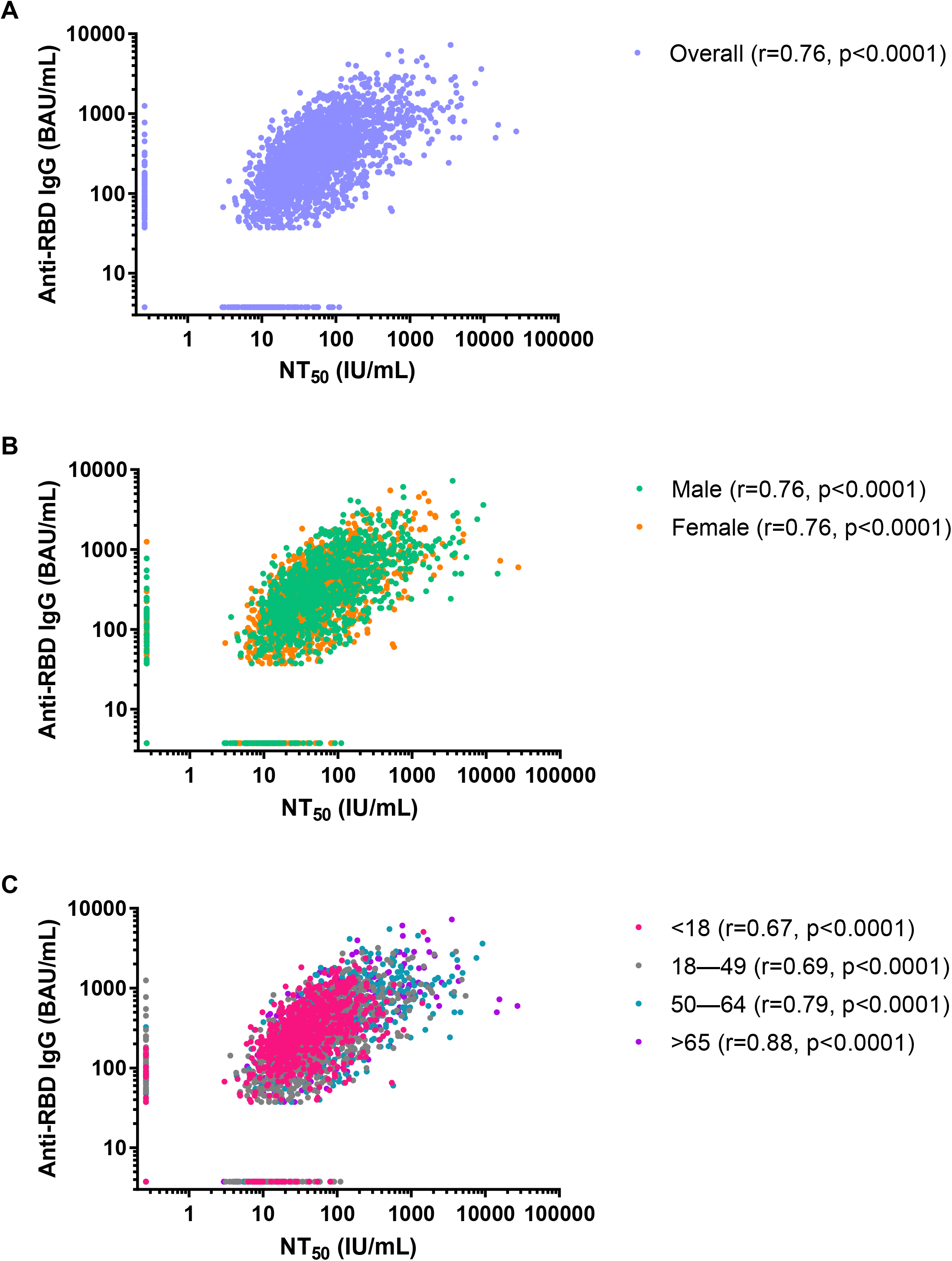
Correlations between SARS-CoV-2 50% neutralizing antibody titer (NT_50_) concentrations in international units per mL (IU/mL) and anti-SARS-CoV-2 receptor binding domain (RBD) IgG concentrations in binding antibody units per mL (BAU/mL) for the convenience sample of 3,067 serum specimens collected during July 27,2020-August 27, 2020 with detectable anti-SARS-CoV-2 antibodies on a qualitative assay. A) Overall, B) by sex, and C) by age class.

NT_50_ concentrations in the overall convenience sample of 3,067 serum specimens collected during July 27, 2020-August 27, 2020 with detectable anti-SARS-CoV-2 antibodies on a qualitative assay were 2.7-fold lower and 8.6-fold lower than concentrations reported from serum specimens collected 28 days post COVID-19 vaccination from ChAdOx1 [11] and mRNA-1273 [12] VE study participants classified as not having COVID-19 during the follow-up period, respectively (Figures 3A and B). Likewise, anti-RBD IgG concentrations in the overall convenience sample were 1.5-fold lower and 24.2-fold lower than concentrations reported from serum specimens collected 28 days post COVID-19 vaccination from ChAdOx1 and mRNA-1273 VE study participants classified as not having COVID-19 during the follow-up period, respectively (Figure 3C and D). Overall, 88.3% of serum specimens in this study met or exceeded the concentration of SARS-CoV-2 neutralizing antibodies associated with 70% ChAdOx1 and mRNA-1273 VE, while 32.9% and 27.1% of sera met or exceeded the concentration of SARS-CoV-2 neutralizing antibodies associated with 90% ChAdOx1 and mRNA-1273 VE, respectively (Figure 1A). For binding antibodies, we found that 63.1% and 86.4% of serum specimens from this study met or exceeded the concentration of anti-RBD IgG antibodies associated with 70% ChAdOx1 and mRNA-1273 VE, respectively, and 1.3% and 13.6% of sera met or exceeded the concentration of anti-RBD IgG antibodies associated with 90% ChAdOx1 and mRNA-1273 VE, respectively (Figure 1B). A greater percentage of males and persons aged ≥50 years met or exceeded the concentrations of SARS-CoV-2 neutralizing antibodies and anti-RBD Ig associated with 70% ChAdOx1 and mRNA-1273 VE than females and persons aged <50 years, respectively.

**Figure 3.**
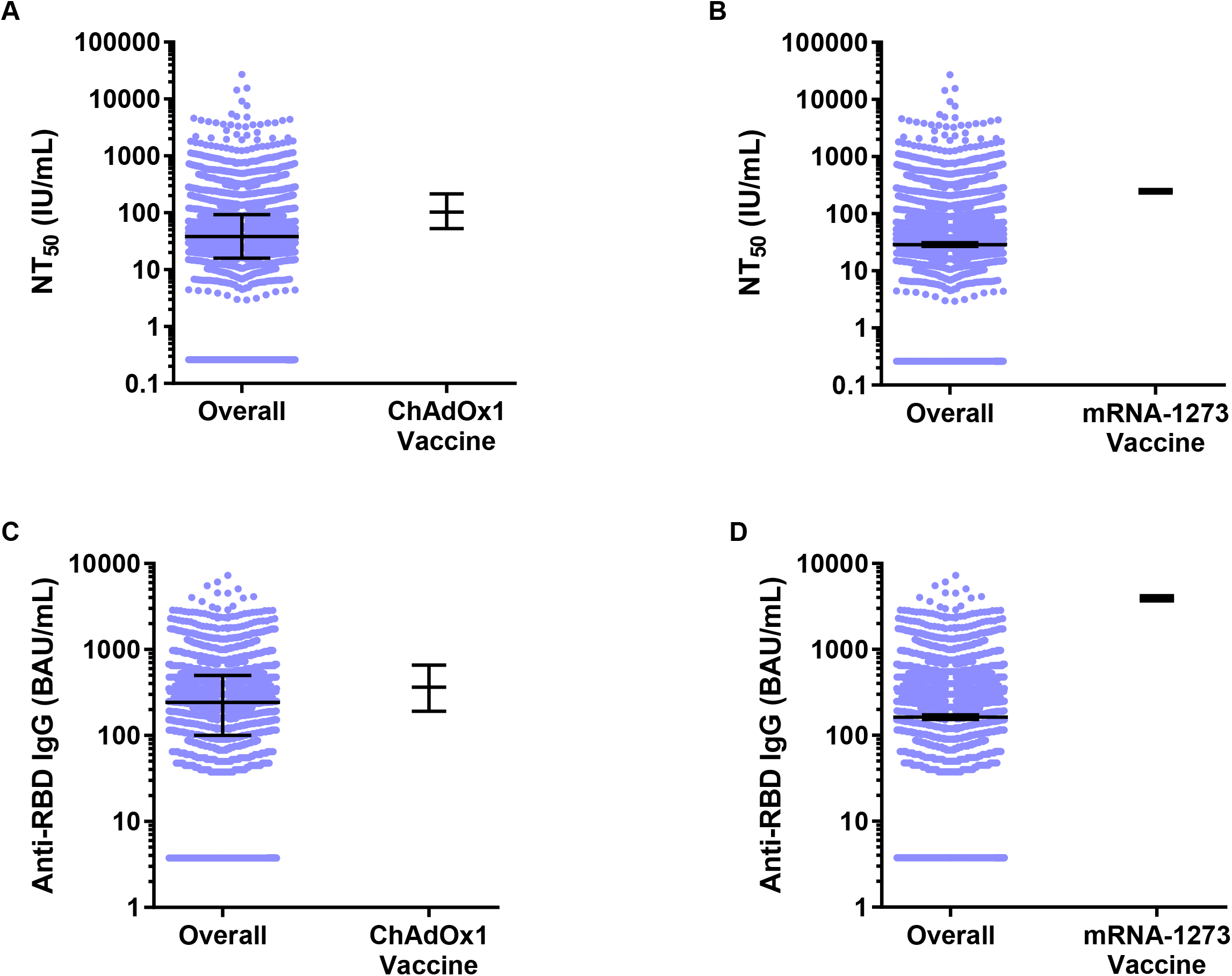
SARS-CoV-2 50% neutralizing antibody titer (NT_50_) concentrations in international units per mL (IU/mL) and anti-SARS-CoV-2 receptor binding domain (RBD) IgG concentrations in binding antibody units per mL (BAU/mL) for the overall convenience sample of 3,067 serum specimens collected during July 27,2020-August 27, 2020 with detectable anti-SARS-CoV-2 antibodies on a qualitative assay compared to 28 days post COVID-19 vaccination concentrations in ChAdOx1 [11] and mRNA-1273 [12] VE study participants classified as not having COVID-19 during the follow-up period. A) Median and interquartile range SARS-CoV-2 NT_50_ concentrations for the overall convenience sample compared to concentrations in VE study participants who received the ChAdOx1 vaccine and were classified as not having COVID-19 during the follow-up period, B) Geometric mean and 95% confidence interval SARS-CoV-2 NT_50_ concentrations for the overall convenience sample compared to concentrations in VE study participants who received the mRNA-1273 vaccine and were classified as not having COVID-19 during the follow-up period, C) Median and interquartile range anti-SARS-CoV-2 RBD IgG concentrations for the overall convenience sample compared to concentrations in VE study participants who received the ChAdOx1 vaccine and were classified as not having COVID-19 during the follow-up period, and D) Geometric mean and 95% CI anti-SARS-CoV-2 RBD IgG concentrations for the overall convenience sample compared to concentrations in VE study participants who received the mRNA-1273 vaccine and were classified as not having COVID-19 during the follow-up period. Horizontal bars represent medians or geometric means, and vertical error bars represent the interquartile range surrounding the median or 95% confidence intervals surrounding the geometric mean.

## DISCUSSION

Similar to the ChAdOx1 [11] and mRNA-1273 [12] VE trial serologic testing results reported in the literature, we demonstrate a statistically significant, strong positive correlation between standardized anti-SARS-CoV-2 RBD IgG and SARS-CoV-2 NT_50_ concentrations, across all sex and age categories, using sera from unvaccinated persons with previous SARS-CoV-2 infection. Based on these broad correlations, our results suggest that high-throughput, quantitative anti-SARS-CoV-2 RBD IgG assays can be used as surrogates for SARS-CoV-2 pseudovirus neutralization assays that are considered mechanistic correlates of protection [12]. Performing high-throughput, commercially available quantitative IgG assays is logistically feasible and practical on a large scale compared to performing time- and resource-intensive neutralization assays. Thus, there is a continued need for FDA-approved quantitative anti-SARS-CoV-2 S and RBD IgG assays in the management of the pandemic to facilitate assessment of antibody concentrations associated with protection from infection and/or disease [14].

While our finding that most serum specimens in this study met or exceeded antibody concentrations associated with 70% COVID VE estimates support the real-world observations of infection-induced immunity from re-infection, it is important to note that less than 33% of sera from unvaccinated persons with previous SARS-CoV-2 infection met or exceeded antibody concentrations associated with 90% ChAdOx1 and mRNA-1273 vaccine efficacy. This suggests that not all persons with a history of SARS-CoV-2 infection generate antibody responses of sufficient magnitude, as measured by a single, randomly timed blood test, to protect them against symptomatic re-infection. In addition to the circulation of the hyper-transmissible Delta variant of SARS-CoV-2, this finding may also partially explain the rise in COVID-19 cases that occurred in the United States in August 2021 despite a national serosurvey in May 2021 showing that 83.3% of the population had SARS-CoV-2 infection- and/or vaccine-induced antibodies [20]. Meanwhile, at least in the short-term (28 days), COVID-19 vaccination can provide 2.7 to 8.6-fold higher neutralizing antibody concentrations and 1.5 to 24.2-fold higher binding antibody concentrations. Thus, our results support current guidance that all eligible persons should consider COVID-19 vaccination to maximize their protection against symptomatic COVID-19 regardless of their SARS-CoV-2 infection history.

Our sampling approach of selecting specimens identified as positive using at least one qualitative anti-SARS-CoV-2 antibody assay would have selected for persons who had more severe disease and thus higher antibody concentrations, and/or in whom antibodies had not waned. This selection bias may have led us to overestimate the percentage of persons with antibody concentrations meeting or exceeding those concentrations associated with 70% and 90% COVID-19 VE and would have missed persons infected with SARS-CoV-2 who did not mount a measurable post-infection immune response or had antibodies below detectable levels.

Our study has several limitations. First, due to the absence of information on whether persons had symptoms or if testing had been done or, if so, the date of COVID-19 symptom onset and/or a positive SARS-CoV-2 qRT-PCR or antigen test, we were unable to calculate the number of days that had elapsed between infection and collection of serum for antibody testing and thus the measurements might not reflect peak antibody concentrations. The ChAdOx1 and mRNA-1273 VE trials measured serum antibody concentrations 28 days after the second vaccine dose, presumably at the peak of the measurable humoral immune response to the vaccine. Therefore, if the sera used in this study were collected >28 days after SARS-CoV-2 infection, it is possible that the percentage of persons with antibody concentrations meeting or exceeding those concentrations associated with 70% and 90% COVID-19 VE were underestimated. However, as a result of the sharp rise in COVID-19 cases in the United States in mid-June 2020 (Supplementary Figure 2) [21], >50% of cases reported in the country prior to July 27, 2020 (date first specimens used in this study were collected) occurred after June 15, 2020. This indicates that the majority of sera in this study were likely to have been collected within 73 days of SARS-CoV-2 infection, a time before IgG antibody concentrations would be expected to have significantly waned from peak post-infection levels [22].

Second, 13.6% of persons had anti-RBD IgG concentrations below the cutoff value of the assay (37.5 BAU/mL). However, the anti-RBD IgG concentration associated with 70% mRNA-1273 VE (8.0 BAU/mL) is below the cutoff value of the Cov2Quant IgG® Assay used in this study. Therefore, it is possible that some persons below the assay cutoff value might have had anti-RBD IgG concentrations greater than or equal to 8.0 BAU/mL, leading us to underestimate the percentage of persons with antibody concentrations meeting or exceeding those concentrations associated with 70% mRNA-1274 COVID-19 VE.

Third, extrapolating these population-level results to individual-level clinical decision making with regard to immune protection based on a single antibody test poses a challenge. The sera tested here were untimed with regard to onset of infection and we have no knowledge of the immune status of the individuals in terms of co-morbidities and medications. These challenges are also applicable to post-vaccination sera in terms of certain populations responding less well to vaccines in terms of neutralizing and binding antibodies as compared to healthy volunteers in vaccine trials and limited availability of standardized quantitative assays [3].

Fourth, our findings, as well as the timing of the ChAdOx1 CoV-19 and mRNA-1273 VE trials, predate the circulation of the Delta variant of SARS-CoV-2 that exhibits a decreased sensitivity to post-vaccination antibody neutralization compared to the Alpha variant of the virus [23]. Therefore, our data is not able to address potential protection of persons infected SARS-CoV-2 in mid-2020 against infection with the Delta variant of the virus.

Finally, the efficiency and redundancy of the immune system, especially for the prevention of severe disease, in terms of humoral [12], cellular immunity, memory and anamnestic response on second exposure to SARS-CoV-2 likely contribute to protection beyond an estimate indicated solely by single antibody concentrations [6, 24].

In conclusion, we demonstrate that in this sample, most non-vaccinated persons with qualitative antibody evidence of prior infection had quantitative antibody concentrations against SARS-CoV-2 that met or exceeded antibody levels associated with 70% VE. However, only a small proportion had antibody responses that met or exceeded levels associated with 90% VE. Our findings suggest that persons with a prior COVID-19 would benefit from vaccination to maximize protective antibody concentrations against symptomatic COVID-19.

## Data Availability

All data produced in the present work are contained in the manuscript.

## Acknowledgments

The authors thank Marla Williams, Morgan Herle, Wade Tanico, and Paul Theobald from Labcorp for help received in planning and execution of sample logistics and data acquisition. A special thanks goes to the reference laboratory operators that performed Cov2Quant IgG® and PhenoSense® CoV Neutralizing Antibody assays. We also thank members of the Multistate Assessment of SARS-CoV-2 Seroprevalence (MASS) steering group, including Lyle Peterson, Margaret Honein, Adam MacNeil, Eduardo Azziz-Baumgartner, Francisco Averhoff, Sridhar Basavaraju, Tina Benoit, Carla Black, Kevin Berney, Ryan Weigand, Michele Owen, Ruchi Pancholy, and Lisa Grohskopf.

## Disclaimer

The findings and conclusions in this report are those of the authors and do not necessarily represent the official position of the Centers for Disease Control and Prevention.

## REFERENCES

1. US Centers for Disease Control and Prevention. COVID-19 Vaccinations in the United States. Available at: https://covid.cdc.gov/covid-data-tracker/#vaccinations_vacc-total-admin-rate-total. Accessed October 14, 2021.

2. Jones JM. About One in Five Americans Remain Vaccine-Resistant. Available at: https://news.gallup.com/poll/353081/one-five-americans-remain-vaccine-resistant.aspx. Accessed September 12, 2021.

3. US Centers for Disease Control and Prevention. Science Brief: SARS-CoV-2 Infection-induced and Vaccine-induced Immunity. Available at: https://www.cdc.gov/coronavirus/2019-ncov/science/science-briefs/vaccine-induced-immunity.html. Accessed November 5, 2021 2021.

4. Chandrashekar A, Liu J, Martinot AJ, et al. SARS-CoV-2 infection protects against rechallenge in rhesus macaques. Science 2020; 369:812–7.

5. McMahan K, Yu J, Mercado NB, et al. Correlates of protection against SARS-CoV-2 in rhesus macaques. Nature 2021; 590:630–4.

6. Corbett KS, Nason MC, Flach B, et al. Immune correlates of protection by mRNA-1273 vaccine against SARS-CoV-2 in nonhuman primates. Science 2021; 373:eabj0299.

7. O’Brien MP, Forleo-Neto E, Musser BJ, et al. Subcutaneous REGEN-COV Antibody Combination to Prevent Covid-19. N Engl J Med 2021; 385:1184–95.

8. Lumley SF, O’Donnell D, Stoesser NE, et al. Antibody Status and Incidence of SARS-CoV-2 Infection in Health Care Workers. N Engl J Med 2021; 384:533–40.

9. Mattiuzzo G, Bentley EM, Hassall M, et al. Establishment of the WHO International Standard and Reference Panel for anti-SARS-CoV-2 antibody (WHO/BS/2020/2403): World Health Organization, 2020.

10. Earle KA, Ambrosino DM, Fiore-Gartland A, et al. Evidence for antibody as a protective correlate for COVID-19 vaccines. Vaccine 2021; 39:4423–8.

11. Feng S, Phillips DJ, White T, et al. Correlates of protection against symptomatic and asymptomatic SARS-CoV-2 infection. medRxiv 2021.

12. Gilbert PB, Montefiori DC, McDermott A, et al. Immune Correlates Analysis of the mRNA-1273 COVID-19 Vaccine Efficacy Trial. medRxiv 2021.

13. Khoury DS, Cromer D, Reynaldi A, et al. Neutralizing antibody levels are highly predictive of immune protection from symptomatic SARS-CoV-2 infection. Nat Med 2021; 27:1205–11.

14. Gundlapalli AV, Salerno RM, Brooks JT, et al. SARS-CoV-2 Serologic Assay Needs for the Next Phase of the US COVID-19 Pandemic Response. Open Forum Infect Dis 2021; 8:ofaa555.

15. Ju B, Zhang Q, Ge J, et al. Human neutralizing antibodies elicited by SARS-CoV-2 infection. Nature 2020; 584:115–9.

16. Robbiani DF, Gaebler C, Muecksch F, et al. Convergent antibody responses to SARS-CoV-2 in convalescent individuals. Nature 2020; 584:437–42.

17. Bajema KL, Wiegand RE, Cuffe K, et al. Estimated SARS-CoV-2 Seroprevalence in the US as of September 2020. JAMA Intern Med 2021; 181:450–60.

18. Markmann AJ, Giallourou N, Bhowmik DR, et al. Sex disparities and neutralizing antibody durability to SARS-CoV-2 infection in convalescent individuals. medRxiv 2021.

19. Kappelman MD, Weaver KN, Boccieri M, et al. Humoral Immune Response to Messenger RNA COVID-19 Vaccines Among Patients With Inflammatory Bowel Disease. Gastroenterology 2021.

20. Jones JM, Stone M, Sulaeman H, et al. Estimated US Infection-and Vaccine-Induced SARS-CoV-2 Seroprevalence Based on Blood Donations, July 2020-May 2021. JAMA 2021.

21. US Centers for Disease Control and Prevention. Trends in Number of COVID-19 Cases and Deaths in the US Reported to CDC, by State/Territory. Available at: https://covid.cdc.gov/covid-data-tracker/#trends_dailycases. Accessed October 14, 2021.

22. Egbert ER, Xiao S, Colantuoni E, et al. Durability of Spike Immunoglobin G Antibodies to SARS-CoV-2 Among Health Care Workers With Prior Infection. JAMA Netw Open 2021; 4:e2123256.

23. Planas D, Veyer D, Baidaliuk A, et al. Reduced sensitivity of SARS-CoV-2 variant Delta to antibody neutralization. Nature 2021; 596:276–80.

24. Sette A, Crotty S. Adaptive immunity to SARS-CoV-2 and COVID-19. Cell 2021; 184:861–80.

