## Supplementary Information for "SARS-CoV-2 Convalescent Sera Binding and Neutralizing Antibody Concentrations Compared with COVID-19 Vaccine Efficacy Estimates Against Symptomatic Infection"

**Supplementary Figure 1.** Frequency distribution for the convenience sample of 3,067 serum specimens collected during July 27,2020-August 27, 2020 with detectable anti-SARS-CoV-2 antibodies on a qualitative assay by jurisdiction.

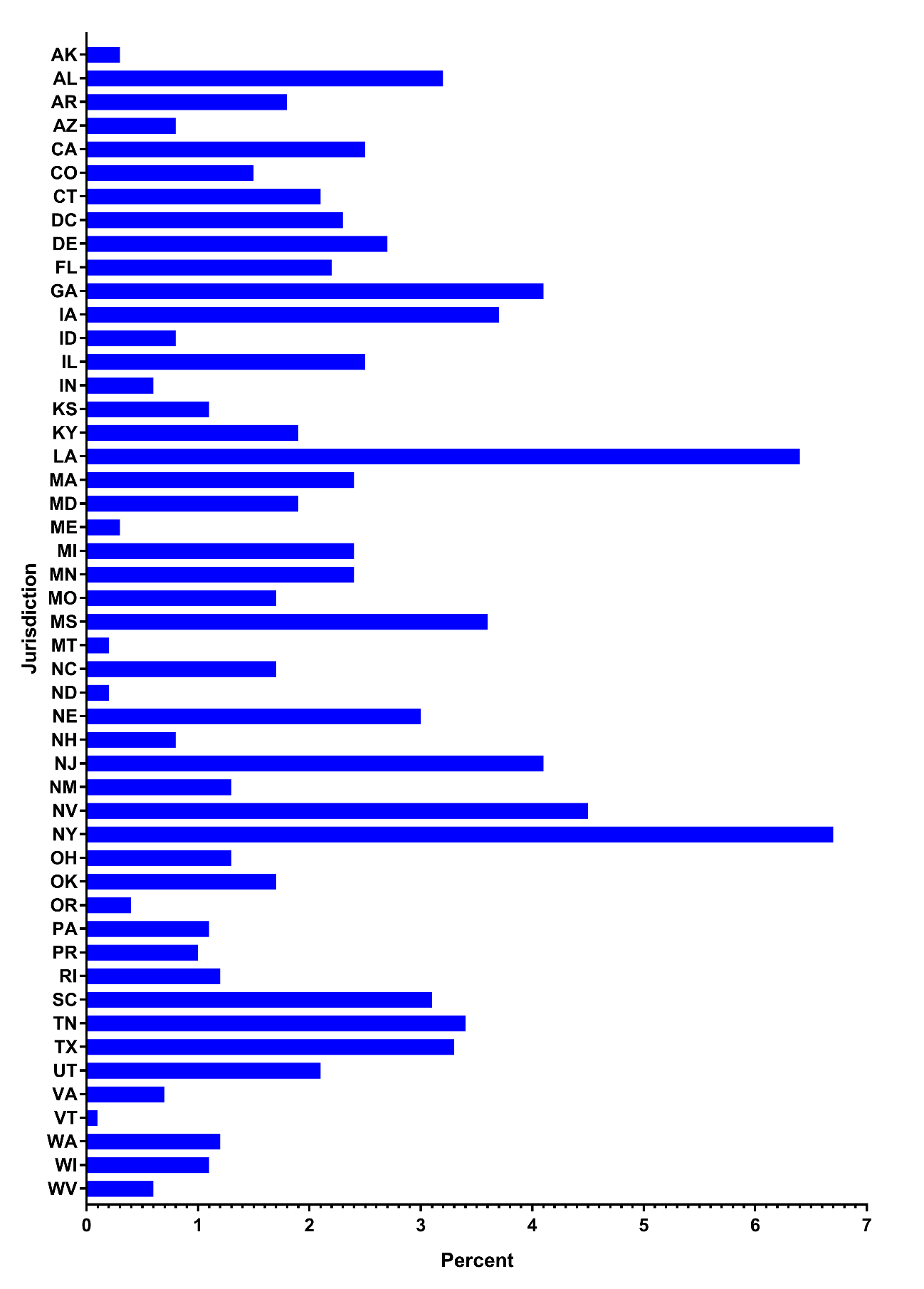

**Supplementary Figure 2.** Number of new SARS-CoV-2 cases in the US (orange; [CDC COVID Data Tracker](https://covid.cdc.gov/covid-data-tracker/#trends_dailycases)) overlaid with the frequency distribution for the convenience sample of 3,067 serum specimens collected during July 27, 2020-August 27, 2020 with detectable anti-SARS-CoV-2 antibodies on a qualitative assay by collection date (blue). As a result of the sharp rise in COVID-19 cases in the United States in mid-June 2020, >50% of cases reported in the country prior to July 27, 2020 (date first specimens tested in this study were collected) occurred after June 15, 2020. This indicates that most sera in this study were likely to have been collected within 73 days of SARS-CoV-2 infection.

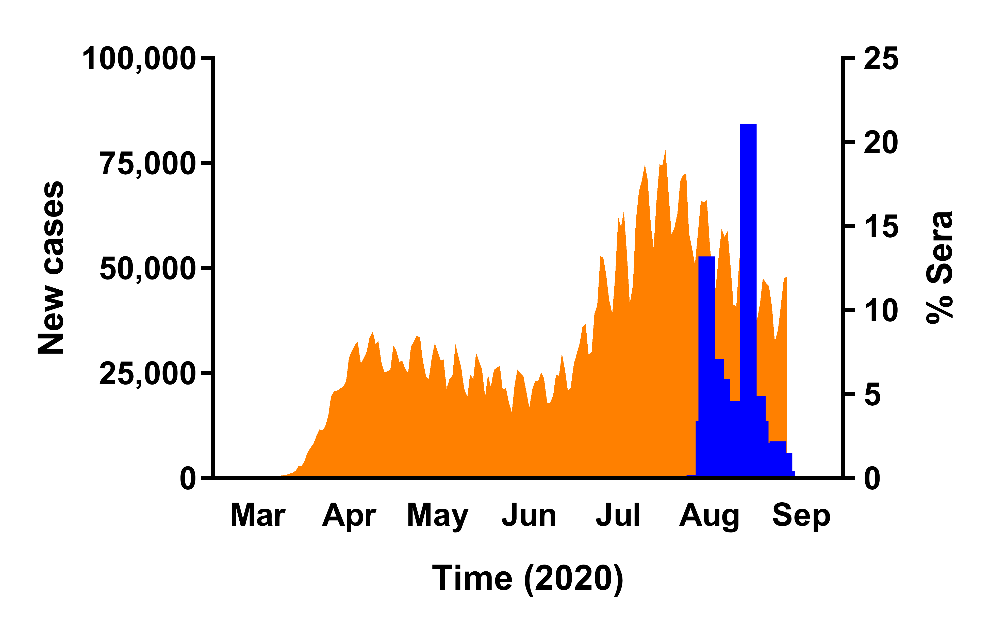

**Supplementary Figure 3.** Frequency distribution for the convenience sample of 3,067 serum specimens collected during July 27, 2020-August 27, 2020 with detectable anti-SARS-CoV-2 antibodies on a qualitative assay by assay type.

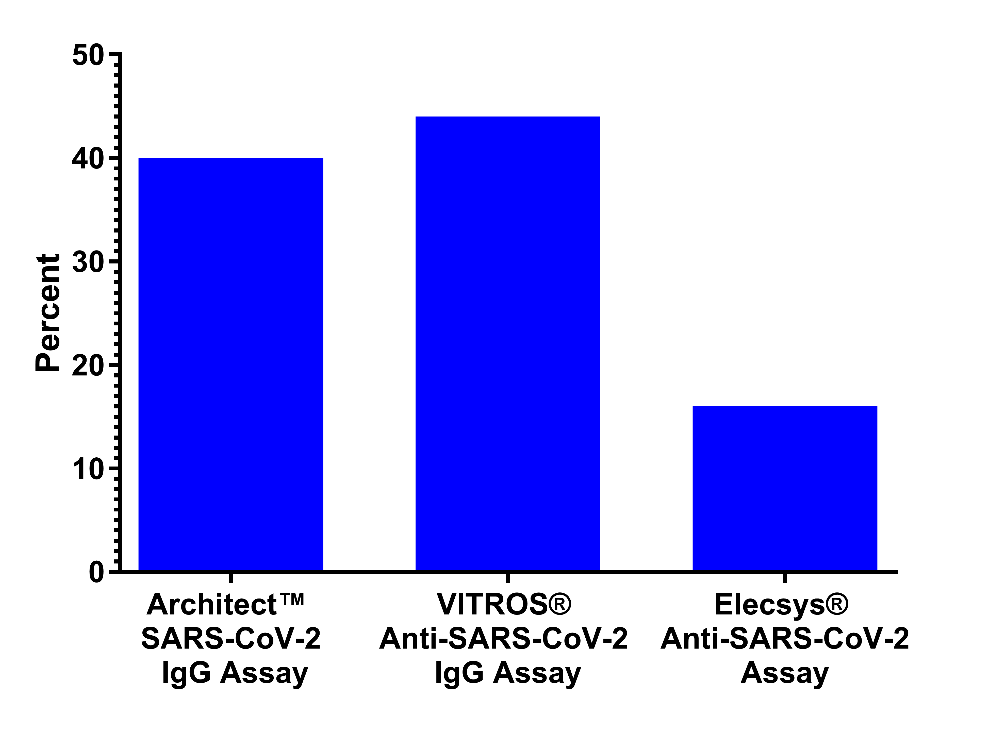

**Supplementary Figure 4.** SARS-CoV-2 50% neutralizing antibody titer (NT_50_) concentrations and anti-SARS-CoV-2 receptor binding domain (RBD) IgG concentrations for the convenience sample of 3,067 serum specimens collected during July 27, 2020-August 27, 2020 with detectable anti-SARS-Cov-2 antibodies on a qualitative assay by assay type. A) SARS-CoV-2 NT_50_ concentrations in international units per mL (IU/mL) and B) Anti-SARS-CoV-2 RBD IgG concentrations in binding antibody units per mL (BAU/mL). Horizontal bars represent geometric means, vertical error bars represent 95% confidence intervals, and dashed horizontal lines represent assay cut-off values for seropositivity. P-values from post-hoc Tukey tests are shown for each qualitative antibody test comparison. Bolded p-values denote statistical significance (p<0.05).

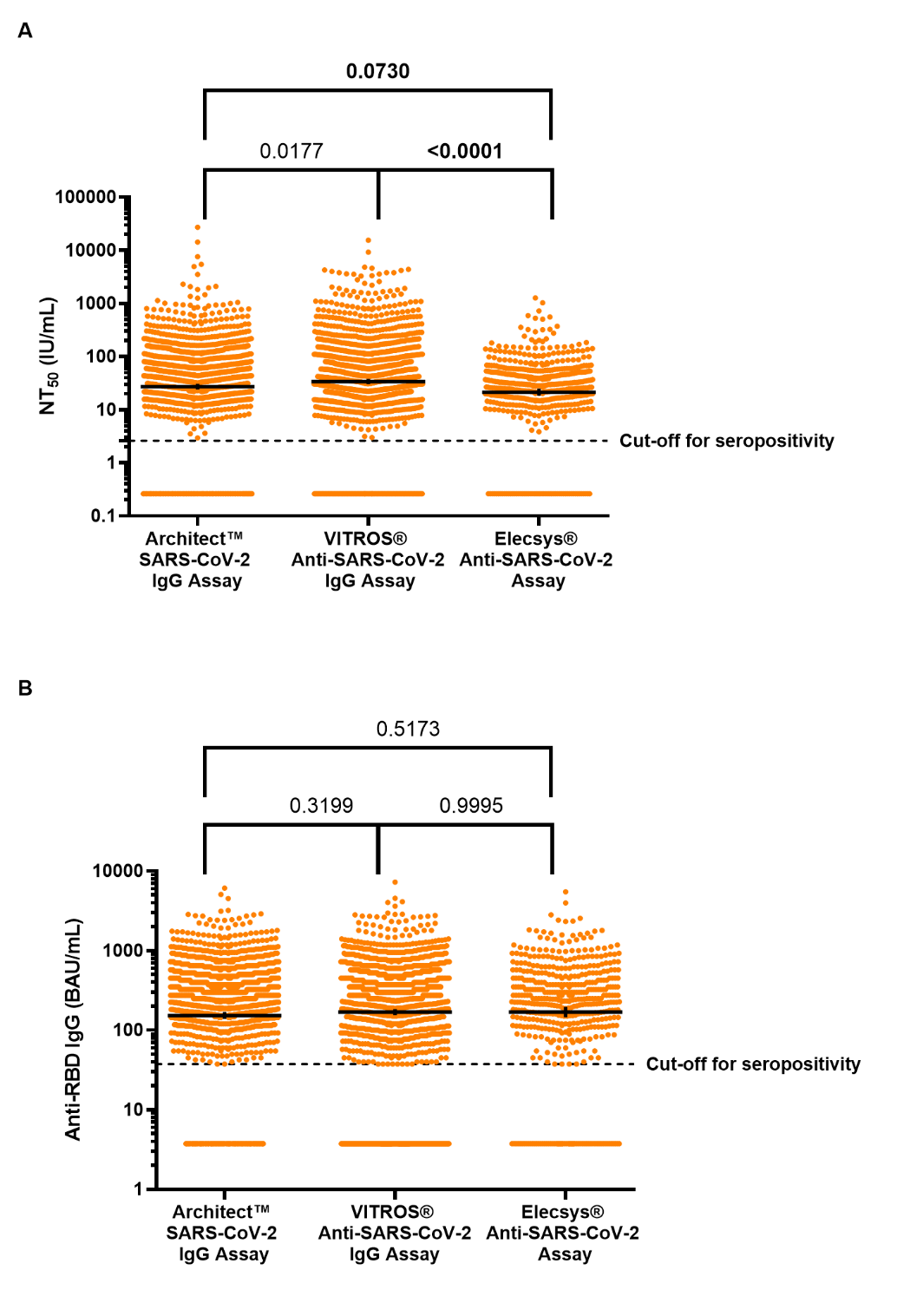
